# The negative acute phase response and not serum C-reactive protein is a major biomarker of major depression: a precision nomothetic psychiatry study

**DOI:** 10.1101/2025.08.15.25333787

**Authors:** Michael Maes, Mengqi Niu, Xu Zhang, Jing Li, Drozdstoy Stoyanov, Bo Zhou, Abbas F. Almulla, Yingqian Zhang

## Abstract

**Background:** Recently, it was suggested that serum high-sensitive C-reactive protein (hsCRP) serves as a biomarker for “inflammatory major depression (MDD)” when serum hsCRP exceeds > 3.0 mg/L. Since 1991-1992, it has been known that MDD is characterized by a negative acute phase protein (APP) response with lowered serum albumin and transferrin levels.

**Aims:** To compare hsCRP with negative APPs as biomarkers of MDD and the severity of physio-affective symptoms and to clarify their relationship with immune and metabolic variables.

**Methods:** This case-control study included 125 MDD patients and 40 healthy controls and assessed serum hsCRP, albumin, transferrin, M1 macrophage profile, and the compensatory immunoregulatory system (CIRS).

**Results:** No significant elevations in hsCRP levels in MDD compared to controls were found. A minority of MDD patients (15.2%) had hsCRP values higher than 3 mg/L, and 78.9% of those had concentrations between 3 and 7 mg/L. Serum hsCRP is largely associated with metabolic parameters, including metabolic syndrome and body mass index (BMI), and with adverse childhood experiences (ACEs), M1 macrophage profile (all positively), and CIRS (inversely). Serum transferrin and albumin are significantly reduced in MDD with an accuracy of 77.3%. Both proteins are strongly and inversely correlated with the physio-affective phenome of MDD, ACEs, and immune activation.

**Conclusions:** MDD is accompanied by non-classical inflammation with a negative APP response associated with immune activation and ACEs. Many MDD patients with hsCRP values <3 mg/L show indicants of a smoldering negative acute phase response. Increased hsCRP does not indicate “inflammatory depression.”

## Introduction

Recent data indicates that major depressive disorder (MDD) is associated with interconnected abnormalities in neuro-immune, metabolic, and oxidative (NIMETOX) pathways [1]. The immunological anomalies during the acute phase of MDD include disruptions in immune homeostasis and a smoldering acute phase response [2–4]. The various stages of MDD—acute, remission, and partial remission—are associated with distinct alterations in homeostasis between the immune-inflammatory response system (IRS) and the compensatory immunoregulatory system (CIRS) [1]. The CIRS controls hyperinflammation, facilitates the return to homeostasis, and possesses healing effects following the resolution of IRS activation [1]. The IRS encompasses various immunological profiles, notably the M1 macrophage profile, characterized by elevated levels of pro-inflammatory cytokines, including interleukin (IL)-6, IL-1, and tumor necrosis factor (TNF)-α and the T helper-17 profile, including IL-17A [3, 5]. Significantly, the M1 cytokines elicit an acute phase response in the liver, marked by diminished plasma concentrations of negative acute phase proteins (APPs), including albumin and transferrin, and elevated levels of positive APPs, such as C-reactive protein (CRP) [1]. Increased IL-17A levels act in synergy with M1 cytokines to enhance inflammation, thereby indirectly contributing to the AP response [6]. The CIRS includes immunoregulatory cytokines such as IL-4, IL-10, and epidermal growth factor (EGF), which collectively can diminish M1 and Th1 polarization [1, 7–9]. Additionally, several immune receptor antagonists, like the soluble IL-1 receptor antagonist (sIL-1RA), indicate IRS and M1 activation while demonstrating immunoregulatory effects [10]. The acute phase of MDD is accompanied by activation of M1, Th-17 and CIRS immune profiles [1].

In the past decade, investigations have suggested that serum high-sensitive CRP (hsCRP) may serve as a biomarker to differentiate “inflammatory depression” or “immune-mediated depression” when serum hsCRP is exceeds > 3.0 mg/L [11]. Certain authors advocate for the stratification of MDD patients based on hsCRP levels at the 3.0 mg/dL threshold to initiate treatment with anti-inflammatory agents, such as anti-TNF and cyclooxygenase-2 (COX-2) inhibitors [11, 12]. Some suggest employing the same hsCRP cut-off value or other immune biomarkers to incorporate “inflammatory depression”, into the upcoming DSM-6 as a distinct phenotype of MDD [13].

Nonetheless, heightened concentrations of hsCRP have been persistently documented in individuals with metabolic disorders, including obesity and metabolic syndrome (MetS) [14–18]. In various investigations in MDD, the correlation between hsCRP and depressive symptoms was no longer evident following adjustments for body mass index (BMI), suggesting that obesity may play a mediating or confounding role in this relationship [19, 20]. Furthermore, it is noteworthy that, in a major depressive episode, nearly 50% of the variance observed in hsCRP levels can be ascribed to a range of factors, including BMI, adverse childhood experiences (ACEs), and age, rather than being directly linked to depression [21].

Elevated levels of hsCRP may indeed signify the existence of an inflammatory process, contingent upon the concentrations observed in serum or plasma. Consequently, values below 3 mg/L are regarded as falling within the normal range [18]. Concentrations ranging from 3 to 10 mg/L suggest normal or slightly elevated levels. The observation of a modest elevation in serum hsCRP concentration, ranging from 3 to 13 mg/L, suggests the presence of metabolic disorders, which may encompass nonalcoholic fatty liver disease, hepatic steatosis, insulin resistance, and preclinical atherosclerosis [18]. Serum hsCRP levels ranging from 13 to 30 mg/L signify a distinct metabolic phenotype marked by immune activation, hepatic dysfunctions (evidenced by elevated alkaline phosphatase and γ-glutamyl transferase), platelet activation, and increased uric acid levels [18]. Furthermore, concentrations exceeding 10 mg/L but less than 100 mg/L indicate moderate inflammation, which may be observed in some autoimmune disorders. Lastly, values surpassing 100 mg/L are typically associated with acute bacterial and some viral infections [18]. It has remained uncertain whether, in MDD, hsCRP levels within the lower concentration ranges serve as indicators of metabolic alterations, MDD, ACEs, and/or M1 macrophage activation.

Furthermore, the assessment of a solitary biomarker, such as hsCRP, fails to encompass the intricacies of the NIMETOX biomarkers in MDD [1]. Research conducted in 1991 showed that a combination of negative APPs, including transferrin and albumin, effectively distinguished MDD patients from controls, achieving a sensitivity of 72% and a specificity of 92% for melancholia [2]. These findings suggest that MDD is marked by a negative acute phase response, disruptions in protein homeostasis, and protein malnutrition, including semistarvation resulting from primary undernutrition due to anorexia and weight loss [2]. Anorexia and weight loss are common symptoms of MDD that may be triggered by an activated M1 macrophage profile [2, 3] and synergistically influenced by various anorectic cytokines, including IL-17A [22]. However, there is a lack of evidence comparing the prevalence or accuracy of the negative APPs versus hsCRP as biomarkers of MDD.

Hence, this study was conducted to compare hsCRP levels with negative APPs as biomarkers of MDD and the severity of depressive subdomains (encompassing affective symptoms, chronic fatigue, and weight loss), and to clarify their relationship with M1 macrophage, IL-17A and CIRS profiles, and metabolic variables, including the prevalence of MetS, BMI and waist circumference.

## Methods

### Participants

This research is a case-control and cross-sectional analysis. A total of 165 participants were enrolled, comprising 125 in the MDD study group and 40 in the healthy control group. Patients were enrolled in the Psychiatric Center of Sichuan Provincial People’s Hospital in Chengdu, China. The criteria for inclusion were as follows: (a) individuals aged 18-65 years of both genders; (b) written informed consent from the patient and their guardian; (c) fulfillment of the diagnostic criteria for MDD as outlined in the Diagnostic and Statistical Manual of Mental Disorders, Fifth Edition (DSM-5); and (d) a Hamilton Depression Scale-21 (HAMD-21) score [23] exceeding 18. The healthy control group comprises staff members, family members of staff, and acquaintances of individuals with MDD. Controls were matched to patients based on age, sex, education, and body mass index. The exclusion criteria for patients and controls included: (a) diagnoses of other significant mental disorders, such as bipolar disorder, schizophrenia, schizoaffective disorder, psycho-organic disorders, substance use disorders (excluding nicotine dependence), and autism spectrum disorders; (b) personality disorders (e.g., borderline, antisocial) and developmental disorders (e.g., severe intellectual disability), as well as neurological conditions like stroke, epilepsy, brain tumors, Parkinson’s disease, Alzheimer’s disease, and multiple sclerosis; (c) major medical conditions, including autoimmune and immunological diseases, psoriasis, systemic lupus erythematosus, inflammatory bowel disease, rheumatoid arthritis, type 1 diabetes mellitus, chronic obstructive pulmonary disease, and cancer. (d) pregnant and lactating women; (e) individuals with a severe allergic reaction in the preceding month; (f) patients with a history of infection in the last three months; (g) patients undergoing treatment with immunosuppressive or immunomodulatory medications, including glucocorticoids; (h) individuals consuming therapeutic doses of antioxidants or omega-3 supplements within the past three months; (i) individuals with a surgical history in the last three months; or (j) frequent analgesic users. Moreover, healthy controls were excluded if they had a history of MDD or dysthymia (lifetime or current), any DSM-IV anxiety disorder, or a familial history of affective disorders, suicide, or substance use disorders (except nicotine dependency).

### Clinical assessments

All participants were interviewed by a qualified researcher (physician) utilizing a semi-structured interview to evaluate demographic and clinical data, including age, gender, educational attainment, income, disease progression, medical history, frequency of depressive episodes, symptoms, personal history, and family history, among others. The Mini International Neuropsychiatric Interview (M.I.N.I.) [24] was utilized to screen for and diagnose major mental disorders and one personality disorder based on the criteria established in the DSM-IV and the International Statistical Classification of Mental Disorders (ICD-10). The scale is intended to assess both current and lifetime psychiatric disorders, which encompass: depressive episodes, dysthymia, hypomanic episodes, suicidal ideation, panic disorder, agoraphobia, social phobia (social anxiety disorder), generalized anxiety disorder, obsessive-compulsive disorder, post-traumatic stress disorder, alcohol dependence/abuse, non-alcoholic substance dependence/abuse, psychotic disorders, anorexia nervosa, bulimia nervosa, and antisocial personality disorder.

On the same day, the same evaluator administered scales to measure the severity of depression, anxiety, and physio-somatic (psychosomatic) symptoms. We utilized the total score from the 21-item HAMD scale to evaluate the severity of depression [23]. We employed the Hamilton Anxiety Rating Scale (HAMA) scale score to assess the severity of anxiety [25]. The State-Trait Anxiety Inventory (STAI), state version, was employed to evaluate self-reported anxiety [26]. The FibroFatigue Scale (FFS), a clinical interview consisting of 12 items, was utilized as a quantitative assessment of symptom intensity for Fibromyalgia and Chronic Fatigue Syndrome (CFS) symptoms [27]. Based on these clinical severity rating scales we constructed an integrated clinical severity index as a z unit-based composite score, namely the sum of z HAMD + z HAMA + z STAI + z FF. The weight loss item of the HAMD was used to assess weight loss in the participants.

The Columbia-Suicide Severity Rating Scale (C-SSRS) [28] was employed to evaluate the cumulative instances of lifetime suicide attempts and suicidal ideation [29]. This study employs two items from the C-SSRS to calculate the recurrence of illness (ROI) index, which is the z unit-based composite score of sum of the z transformations of number of depressive episodes, the number of lifetime suicidal attempts (up to one month before the index episode), and the number of suicidal ideations (up to one month before the index episode) [29]. We employed the Childhood Trauma Questionnaire-Short Form (CTQ-SF) to evaluate the severity of Adverse Childhood Experiences (ACEs) [30]. Zhao Xingfu translated the questionnaire’s Chinese version [31]. As previously detailed, we calculated the scores across the five subscales: emotional abuse, physical abuse, sexual abuse, emotional neglect, and physical neglect [32]. This study utilized the aggregate of the five subscale scores to quantify the severity of all ACEs.

Body weight, height, heart rate, and blood pressure were evaluated. BMI was calculated as weigh in kilogram divided on height in meters squared. The waist circumference (WC), a measurement that is employed to estimate the amount of subcutaneous abdominal fat, was measured in the horizontal plane, located halfway between the iliac crest and the lowest ribcage. As an integrated index of BMI and WC we computed a z compote score as z BMI + z WC. MetS is characterized by the presence of three or more of the following components, as delineated in the 2009 Joint Scientific Statement issued by the American Heart Association and the National Heart, Lung, and Blood Institute [33]: (a) A waist circumference of 90 cm or greater for males and 80 cm for females; (b) A triglyceride level of 150 mg/dL or higher; (c) A HDL cholesterol level of less than 40 mg/dL for males and less than 50 mg/dL for females; (d) Elevated blood pressure defined as greater than 130 mm Hg systolic or 85 mm Hg diastolic, or the use of antihypertensive medication; (e) An increase in fasting glucose levels of 100 mg/dL or more, or a diagnosis of diabetes. Based on the number of MetS criteria we used the MetS ranking in the analyses.

Assays.

A total of 30 milliliters of fasting venous blood were obtained from each participant between 6.30 and 8:00 a.m. using serum tubing and a disposable syringe. After centrifugation of blood at 3500 rpm, serum was collected and preserved in Eppendorf tubes with small aliquots at - 80 °C until thawed for biomarker testing. Serum hsCRP was determined using a particle-enhanced immunoturbidimetric assay (DIAYS DIAGNOSTIC SYSTEM (SHANGHAI) CO., LTD) on a fully automated biochemical analyzer (ADVIA 2400, Siemens Healthcare Diagnostics Inc) with a sensitivity of 0.3 mg/L, intra-assay, and inter-assay analytical CVs of 2.80 and 2.17%, respectively. Serum transferrin was measured using an immunoturbidimetric assay (DIAYS DIAGNOSTIC SYSTEM (SHANGHAI) CO., LTD) on a fully automated biochemical analyzer (ADVIA 2400, Siemens Healthcare Diagnostics Inc) with a sensitivity of 0.03 g/L, intra-assay, and inter-assay analytical coefficients of variation (CVs) of 1.96% and 0.67%, respectively. Serum albumin was detected by the Bromocresol Green Method kit (Beijing Strong Biotechnologies, Inc.) on a fully automated biochemical analyzer (ADVIA 2400, Siemens Healthcare Diagnostics Inc.) with the intra-assay and inter-assay analytical CVs of 1.20% and 2.10%, respectively. To compute an integrated index of the negative AP response, we computed a z unit-based composite score as z albumin + z transferrin.

Serum cytokines were assayed using the Luminex xMAP technology (Luminex Corporation, Austin, TX, USA) on the Luminex 200 system. The Human XL Cytokine Fixed Panel (Cat. No.: LKTM014B, bio-techne, R&D Systems) enabled us to measure the fluorescence intensity (FI) and concentrations of 46 cytokines, chemokines, and growth factors in the serum. The blank was subtracted from the IF to obtain accurate results. Briefly, the procedure consisted of the following steps: 1) Made a 2-fold dilution with Calibrator Diluent RD6-65; 2) Added the diluted samples (50 μL) and the diluted microparticle cocktail (50 μL) per well to the 96-well plate, incubated at room temperature with shaking at 850 rpm for 2 hours; 3) Washed the plate three times, added 50 μL of diluted Biotin-Antibody Cocktail each well and incubated for 1 hour at room temperature on the shaker set at 850 rpm; 4) Repeated the wash as the step above, and added 50 μL of diluted Streptavidin-PE to each well. Incubated for 30 minutes at room temperature (850 rpm); 5) Repeated the wash step, resuspended the microparticles by adding 100 μL of wash buffer for 2 minutes in each well. The subsequent analysis employed the Luminex 200 System to assess the 46 cytokines. For all analytes, the intra-assay CVs were lower than 5%, and inter-assay CVs were lower than 11.2%. In the current study, we used the levels of IL-1β, IL-6, TNF-α and sIL-1RA to compute a z unit-based composite score reflecting M1 macrophage activity [5]. CIRS activity was conceptualized as a z unit-based composite score based on measurements of IL-4, IL-10, and EGF [5]. We also used IL-17A as an indicant of Th-17 activity [5]. A comprehensive analysis of all immune profiles in relationship to phenome features will be published separately (Mengqi et al., in preparation).

### Statistics

Pearson’s product-moment and point-biserial correlations were employed to analyze relationships among continuous variables and between continuous and binary data, respectively. Contingency table analysis was employed to investigate the relationship between categorical variables. Analysis of variance (ANOVA) was employed to compare continuous variables across study groups. Numerous comparisons and relationships were adjusted for the False Discovery Rate (FDR). To investigate the distinctions between MDD and control subjects, as well as between individuals with elevated vs diminished hsCRP levels, we performed a binary logistic regression analysis, using MDD or the high CRP group as dependent variables and controls or the low CRP groups as reference categories, respectively. This investigation considered potential confounding variables including age, gender, smoking status, and BMI. Key results encompassed effect size estimates utilizing Nagelkerke pseudo-R square, Wald statistics accompanied by p-values, Odds ratios with 95% confidence intervals (CI), and unstandardized regression coefficient B along with its standard error (SE), where the Wald statistic is defined as the square of the ratio of the B coefficient to its SE. Furthermore, we employed the oversampling technique to compensate for the low prevalence of subgroups, specifically the high hsCRP subgroup in relation to the low hsCRP group, as well as the normal controls group versus the MDD group. We computed the cross-validated accuracy using linear discriminant analysis with 10-fold cross-validation. The area under the receiving operating curve (ROC) was computed together with the Gini-index, Max K-S values, and the model quality. We employed multiple regression analysis to predict acute phase reactants or rating scale scores, using demographic characteristics such as age, gender, metabolic variables, and other biomarkers as explanatory variables. We employed both a manual and an automatic stepwise methodology. The automated approach involved automatic linear modeling with overfitting mitigation, where the criteria for variable inclusion and exclusion were established at p-values of 0.05 and 0.07, respectively. This investigation included model metrics, encompassing standardized beta coefficients, degrees of freedom (df), p-values, R², and F-statistics. We employed the White test and the modified Breusch-Pagan test to investigate heteroskedasticity, whereas collinearity was assessed by tolerance and the variance inflation factor. All tests employed a two-tailed design, with a significance threshold set at 0.05. We utilized IBM’s SPSS 30, Windows edition, for all statistical analyses in this work. In order to normalize the data, we used different transformations where needed, including log10, square root, rank-order, and Winsoring.

Partial least squares (PLS) SEM analysis was employed to examine the causative associations among sex, ACEs, metabolic variables, ROI, immune profiles, albumin, transferrin, hsCRP, and the phenome of depression. The depression phenome, ACEs, and IRS/CIRS (all M1 cytokines and IL-17A, and the CIRS cytokines) were inputted as latent vectors, whereas all other variables were presented as single indicators. One latent vector could be extracted from physical and emotional abuse/neglect, whereas sexual abuse loaded not significantly on this factor and, consequently, was excluded. PLS analysis was conducted solely when the outer and inner models met the pre-specified quality criteria: a) all outer loadings are greater than 0.65 at p < 0.001; b) the latent vectors exhibits robust construct and convergence validity, evidenced by an average variance extracted (AVE) exceeding 0.5, Cronbach’s alpha surpassing 0.7, composite reliability exceeding 0.8, and rho A exceeding 0.8; c) Confirmatory Tetrad Analysis (CTA) confirms that the model is correctly specified as a reflective model; d) discriminatory validity is achieved; and e) the overall model fit, indicated by the standardized root mean square residual (SRMR), is acceptable with a value below 0.08. Upon the fulfillment of the aforementioned model quality data criteria, we perform a PLS-SEM path analysis utilizing 5,000 bootstrap samples, generating path coefficients (accompanied by p-values), and further calculating both specific and total indirect (mediated) effects, along with the total effects.

## Results

### Socio-demographic data

**Table 1** shows the sociodemographic and clinical data of the patients and controls in the current study. There were no significant differences in age, sex ratio, and smoking among MDD patients and controls. There were no significant differences in metabolic variables, including MetS prevalence, MetS ranking, waist circumference and BMI. The total HAMD, HAMA, FF, and STAI scores were significantly higher in MDD as compared with controls. In addition, weight loss scores were significantly higher in patients than controls. There were no significant differences in serum hsCRP and M1 macrophage profiles scores among MDD and controls. Serum albumin and transferrin were significantly lower, whilst IL-17A and the CIRS profile were significantly higher in patients than controls.

**Table 1.**
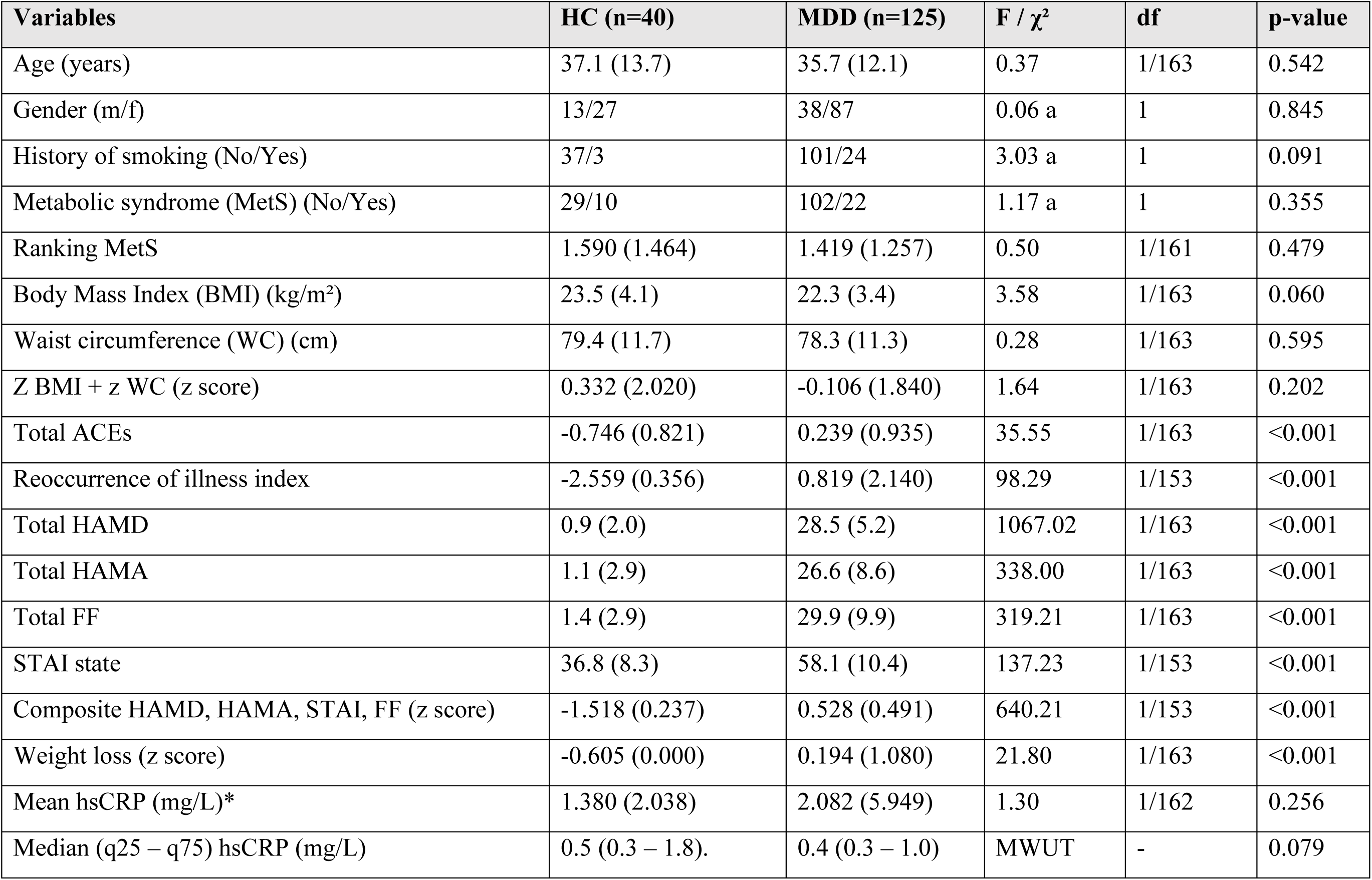
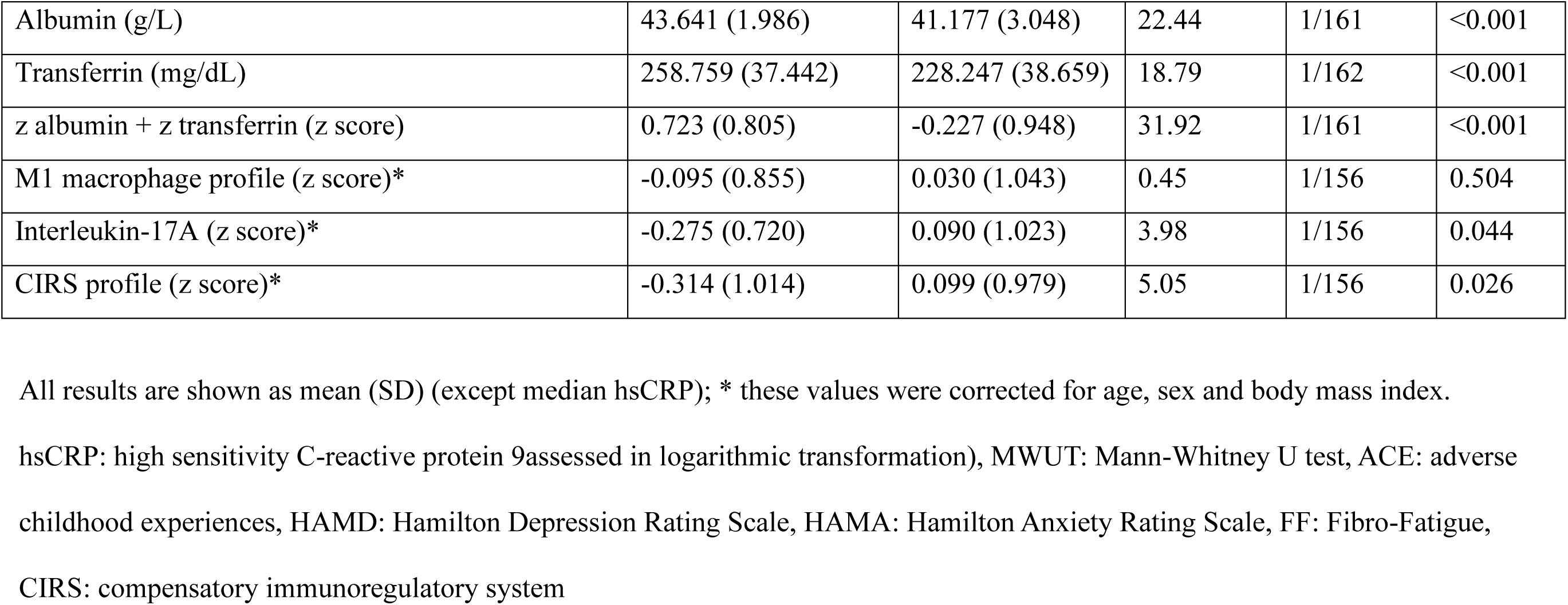
Socio-demographic and clinical data of patients with major depressive disorder (MDD) and healthy controls (HC).

### Features of MDD with and without MetS

**Table 2** shows two different models predicting MDD as dependent variable and controls as the reference group (using binary logistic regression). Model #1 shows that MDD was significantly associated with total ACEs, IL-17A, male sex (positively) and transferrin, albumin and hsCRP (all three inversely) (χ2=123.08, df=6, p<0.001; Nagelkerke=0.545). The accuracy of this model was 79.1% (sensitivity=76.7% and specificity=81.6%). Model #2 examines the impact of both negative acute phase proteins on MDD, indicating that both transferrin and albumin were together significant predictors of MDD (χ2=62.19, df=2, p<0.001; Nagelkerke=0.303). The accuracy of this model was 72.6% (sensitivity=61.3% and specificity=86.6%). The cross-validated accuracy was 69.3% (AUC=0.778, Gini index=0.556, Max K-S=0.483). Adding IL-17A and CIRS shows a cross-validated accuracy of 73.9% (AUC=0.798, Gini index=0.596, Max K-S=0.41). Adding total ACEs yielded a cross-validated accuracy of 77.4% (AUC=0.866, Gini index=0.732, Max K-S=0.648).

**Table 2:**
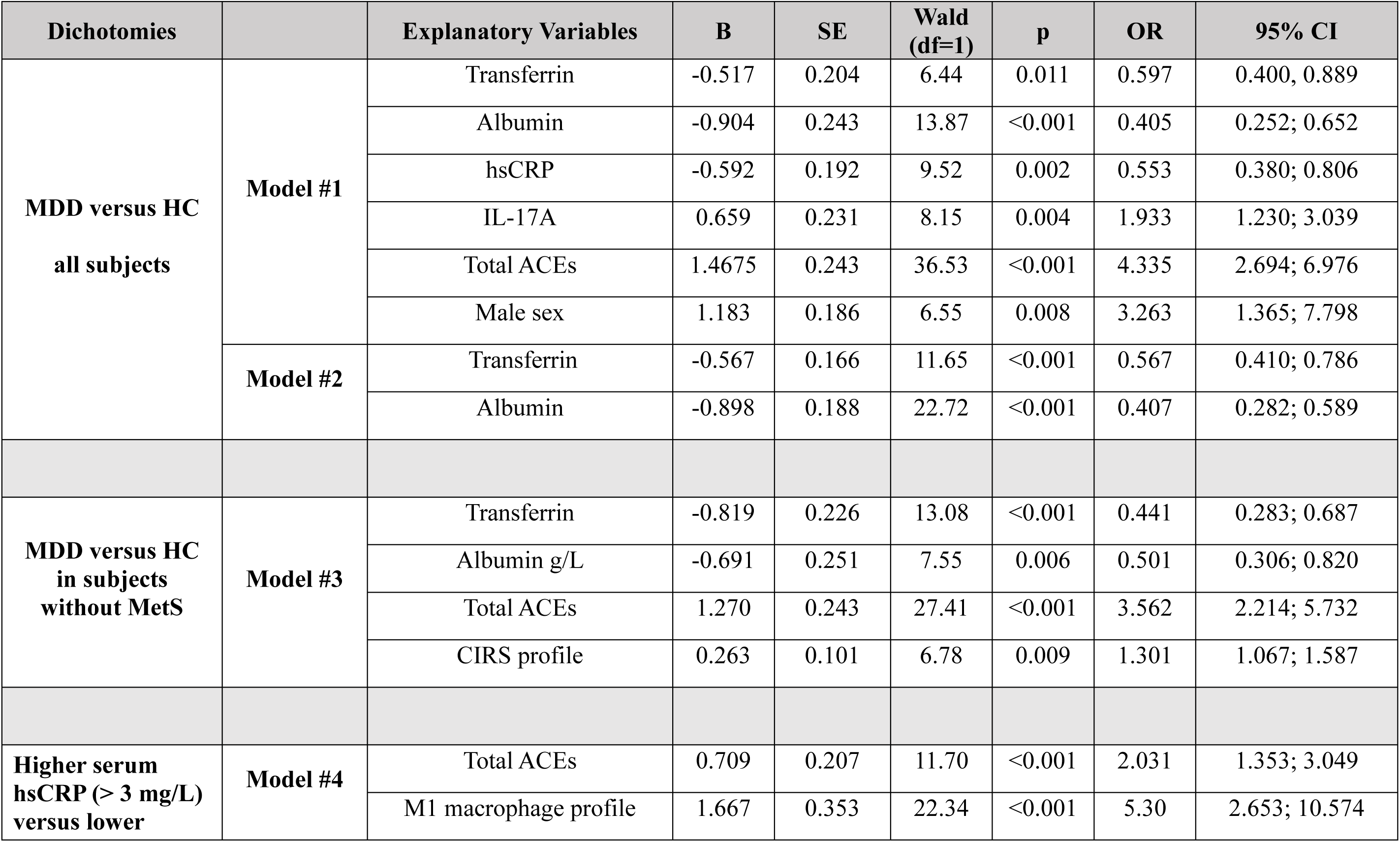
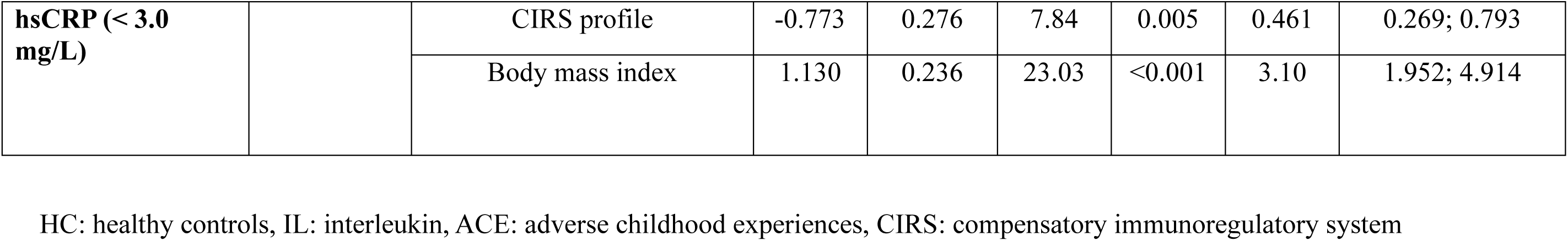
Results of binary logistic regression analysis with major depression (MDD) or increased high sensitivity C-reactive protein (hsCRP) or as dependent variables.

We have also examined the differences between MDD patients and controls after removing subjects with MetS. The results of this binary regression analysis are shown in **Table 3**, model #3. We found that MDD (without MetS) was significantly predicted by lowered transferrin and albumin, and by increased total ACEs and the CIRS profile (χ2=94.18, df=4, p<0.001; Nagelkerke=0.533). The accuracy of this model was 77.3% (sensitivity=76.2% and specificity=78.6%).

**Table 3.** Socio-demographic and clinical data of subjects with low versus high sensitivity C-Reactive Protein (hsCRP) concentrations.

| Variables | hsCRP < 3 (n=142) | hsCRP > 3 (n=22) | F / $\chi^2$ | df | p-value |
| --- | --- | --- | --- | --- | --- |
| Median (q25 – q 75) hsCRP (mg/L) | 0.4 (0.3 – 0.8) | 4.8 (3.78 – 9.18) | MWUT | - | <0.001 |
| Controls / MDD | 36 / 106 | 3 / 19 | 1.44 | 1 | 0.230 |
| Age (years) | 36.2 (12.8) | 33.9 (9.7) | 0.68 | 1/162 | 0.410 |
| Gender (M/F) | 43/99 | 8/14 | 0.33 a | 1 | 0.623 |
| History of smoking (No/Yes) | 120/22 | 17/5 | 0.73 a | 1 | 0.536 |
| Metabolic syndrome (MetS) (No/Yes) | 118/23 | 9/13 | 7.30 a | 1 | 0.011 |
| Rank Mets | 1.35 (1.25) | 2.18 (1.47) | 8.10 | 1/161 | 0.005 |
| Body Mass Index (BMI) (kg/m<sup>2</sup>) | 22.2 (3.5) | 25.1 (3.1) | 14.12 | 1/162 | <0.001 |
| Waist circumference (WC) (cm) | 77.4 (11.3) | 85.9 (9.9) | 11.15 | 1/162 | 0.001 |
| z WC + z BMI (z score) | -0.216 (1.851) | 1.355 (1.607) | 14.18 | 1/162 | <0.001 |
| Total ACEs (z score) | -0.061 (0.995) | 0.375 (0.988) | 3.66 | 1/162 | 0.057 |
| Reoccurrence of illness index | -0.062 (2.408) | 0.141 (2.136) | 0.13 | 1/152 | 0.722 |
| Total HAMD | 21.5 (12.9) | 25.0 (11.4) | 1.47 | 1/162 | 0.227 |
| Total HAMA | 20.0 (13.3) | 23.818 (13.0) | 1.56 | 1/162 | 0.213 |
| Total FF | 22.535 (15.1) | 26.7 (14.5) | 1.49 | 1/162 | 0.225 |
| Total STAI, state | 52.1 (13.9) | 56.6 (12.0) | 1.86 | 1/152 | 0.174 |
| PC HAMD,HAMA,STAI, FF | -0.033 (1.004) | 0.286 (0.931) | 1.79 | 1/152 | 0.183 |
| Weight loss score (z score) | 0.010 (0.995) | -0.038 (1.071) | 0.04 | 1/162 | 0.835 |
| Albumin (g/L) | 41.82 (3.09) | 41.41 (2.48) | 0.37 | 1/161 | 0.546 |
| Transferrin (mg/dL) | 236.1 (41.8) | 231.4 (30.9) | 0.26 | 1/162 | 0.612 |
| z Albumin + z Transferrin (z score) | 0.022 (1.023) | -0.140 (0.848) | 0.49 | 1/161 | 0.483 |
| M1 macrophage profile (z score)* | -0.125 (0.897) | 0.816 (1.255) | 17.86 | 1/156 | <0.001 |
| Interleukin-17A (z score)* | 0.036 (1.013) | -0.238 (0.744) | 1.42 | 1/156 | 0.235 |
| CIRS profile (z score)* | -0.025 (0.994) | 0.163 (1.047) | 0.64 | 1/156 | 0.424 |

All results are shown as mean (SD) (except hsCRP), MWUT: Mann-Whitney U test, \* these values were corrected for age, sex and body mass index.
ACE: adverse childhood experiences, HAMD: Hamilton Depression Rating Scale, HAMA: Hamilton Anxiety Rating Scale, FF: Fibro-Fatigue, CIRS: compensatory immunoregulatory system

### Features of increased hsCRP values

**Table 3** shows the features of subjects with increased hsCRP values. Only 22 participants showed hsCRP values exceeding the threshold value (3 mg/L), namely 19 patients with MDD and 3 controls. There were no significant differences in the control/MDD ratio among patients with higher versus lower hsCRP values. The latter groups did not differ in age, sex ratio, and smoking behavior. All metabolic variables were significantly higher in subjects with higher hsCRP values than in those with lower values, including MetS prevalence, MetS ranking, BMI and waist circumference. There were no significant differences between both groups in total ACEs, ROI, rating scale scores, and weight loss. There were no significant differences in both groups in albumin, transferrin, IL-17A and the CIRS profile. The M1 macrophage profile was significantly higher in people with increased hsCRP as compared with those with a lower hsCRP.

**Table 2, model #4** shows the results of binary logistic regression analysis (oversampling method) with the high CRP group as dependent variable. We found that the latter subgroup was significantly predicted by increased M1 macrophage profile, BMI, and total ACEs (all positively) and the CIRS profile (inversely) (χ2=81.94, df=4, p<0.001; Nagelkerke=0.472). The accuracy of this model was 82.0% (sensitivity=61.9% and specificity=91.2%).

**Table 4** shows the Pearson’s product moment correlations between hcCRP, albumin and transferrin and the z albumin + z transferrin index, and the clinical and immunological variables measured in our study. We found no significant correlations between hsCRP and clinical rating scale scores, total ACEs and ROI. hsCRP was significantly and positively correlated with BMI, waist circumference, rank of MetS, and the M1 macrophage profile.

**Table 4.**
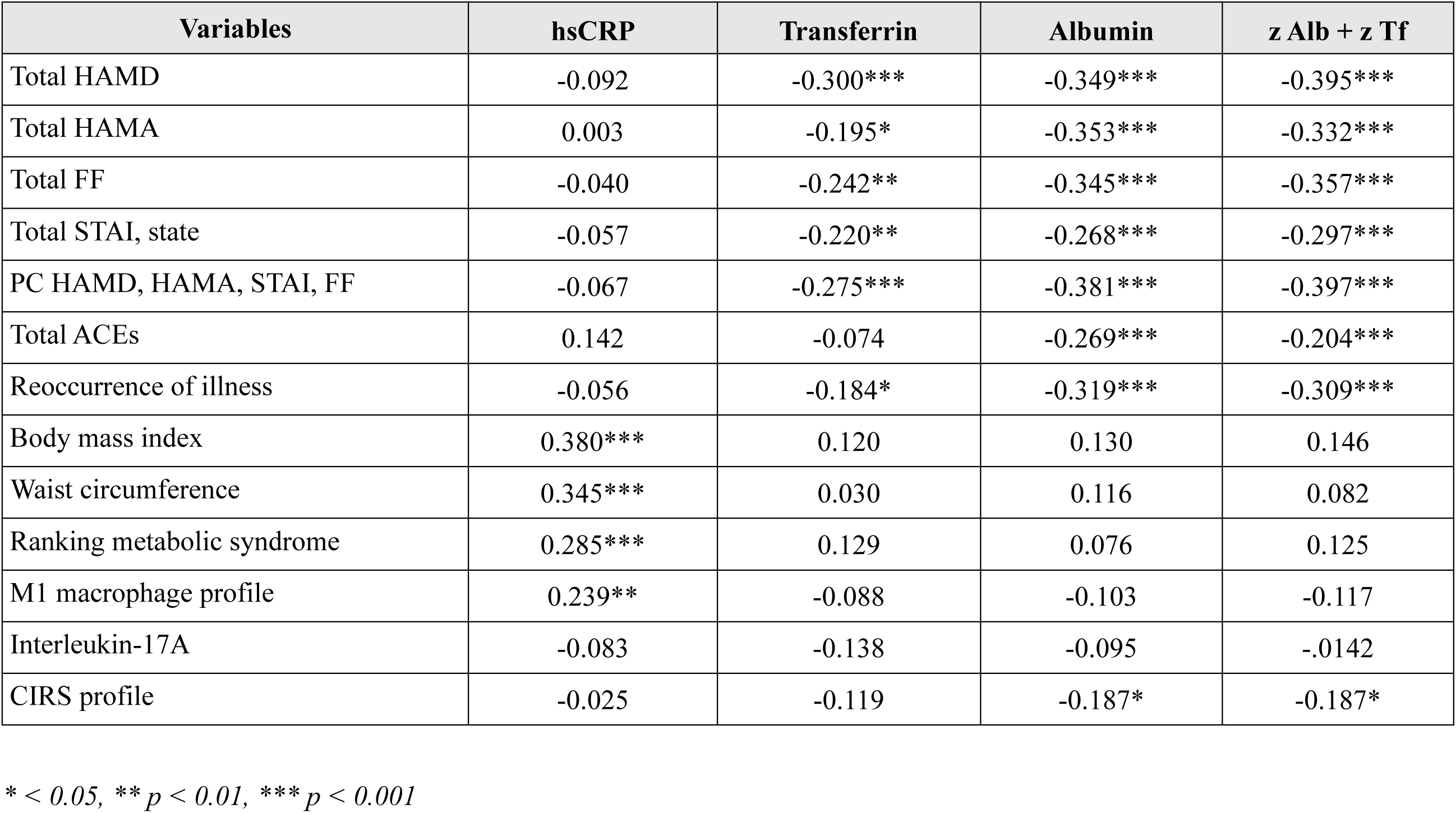
Intercorrelation matrix (Pearson’s correlation coefficients) between biomarkers and severity scales.

| Variables | hsCRP | Transferrin | Albumin | z Alb + z Tf |
| --- | --- | --- | --- | --- |
| Total HAMD | -0.092 | -0.300*** | -0.349*** | -0.395*** |
| Total HAMA | 0.003 | -0.195* | -0.353*** | -0.332*** |
| Total FF | -0.040 | -0.242** | -0.345*** | -0.357*** |
| Total STAI, state | -0.057 | -0.220** | -0.268*** | -0.297*** |
| PC HAMD, HAMA, STAI, FF | -0.067 | -0.275*** | -0.381*** | -0.397*** |
| Total ACEs | 0.142 | -0.074 | -0.269*** | -0.204*** |
| Reoccurrence of illness | -0.056 | -0.184* | -0.319*** | -0.309*** |
| Body mass index | 0.380*** | 0.120 | 0.130 | 0.146 |
| Waist circumference | 0.345*** | 0.030 | 0.116 | 0.082 |
| Ranking metabolic syndrome | 0.285*** | 0.129 | 0.076 | 0.125 |
| M1 macrophage profile | 0.239** | -0.088 | -0.103 | -0.117 |
| Interleukin-17A | -0.083 | -0.138 | -0.095 | -.0142 |
| CIRS profile | -0.025 | -0.119 | -0.187* | -0.187* |

\* < 0.05, \*\* $p < 0.01$ , \*\*\* $p < 0.001$

**Table 5** shows the results of multiple regression analysis with hsCRP as dependent variables and metabolic, immunological, and demographic data as explanatory variables. We examined the prediction of hsCRP in three different groups, namely the total study group, in subjects with a lower hsCRP value (< 3 mg/L) and in those with a higher hsCRP value (> 3.0 mg/L). Model #1 shows that 24.6% of the variance in hsCRP was explained by the regression on BMI, total ACEs, and M1 macrophage profile (all three positively associated and the CIRS profile (inversely associated). In people with a hsCRP values < 3.0 mg/L (model #2), 17.9% of the variance in hsCRP was explained by the z BMI + z WC index and female gender (positively associated). In those with hsCRP values > 3 mg/L, we found that 28.7% of the variance in hsCRP was explained by the rank of MetS.

**Table 5.** Results of multiple regression analysis with the biomarkers as dependent variables and clinical indices as explanatory variables.

| Dependent Variables | Explanatory Variables | Coefficient statistics |  |  | Model statistics |  |  |  |
| --- | --- | --- | --- | --- | --- | --- | --- | --- |
| | | $\beta$ | t | p | R<sup>2</sup> | F | df | p |
| #1. hsCRP in all subjects | **Model** |  |  |  | 0.246 | 12.45 | 4/153 | <0.001 |
|  | Body mass index (BMI) | 0.339 | 4.67 | <0.001 |  |  |  |  |
|  | Total ACEs | 0.201 | 2.83 | 0.005 |  |  |  |  |
|  | M1 macrophage profile | 0.348 | 3.71 | <0.001 |  |  |  |  |
|  | CIRS profile | -0.248 | -2.68 | 0.008 |  |  |  |  |
| #2. hsCRP (in those with CRP < 3.0 mg/L) | **Model** |  |  |  | 0.179 | 14.57 | 2/134 | <0.001 |
|  | z BMI + z WC index | 0.424 | 5.19 | <0.001 |  |  |  |  |
|  | Gender | -0.236 | -2.90 | 0.004 |  |  |  |  |
| #3. hsCRP (in those with hsCRP3 > 3 mg/L) | **Model** |  |  |  | 0.287 | 7.66 | 1/19 | 0.012 |
|  | Ranking Metabolic syndrome | -0.536 | -2.77 | 0.012 |  |  |  |  |
| #4. Albumin in all subjects | **Model** |  |  |  | 0.201 | 9.04 | 4/144 | <0.001 |
|  | Gender | 0.251 | 3.20 | 0.002 |  |  |  |  |
|  | Weight loss score | -0.156 | -1.98 | 0.050 |  |  |  |  |
|  | Total ACEs | -0.187 | -2.38 | 0.019 |  |  |  |  |
|  | CIRS profile | -0.176 | -2.25 | 0.026 |  |  |  |  |
| #5. Albumin in all subjects | **Model** |  |  |  | 0.218 | 13.46 | 3/145 | <0.001 |
|  | Reoccurrence of illness | -0.280 | -3.75 | <0.001 |  |  |  |  |
|  | Gender | 0.271 | 3.65 | <0.001 |  |  |  |  |
|  | Weight loss score | -0.199 | -2.70 | 0.008 |  |  |  |  |
| #6. Transferrin in all subjects | **Model** |  |  |  | 0.194 | 6.87 | 5/143 | <0.001 |
|  | Weight loss score | -0.178 | -2.30 | 0.023 |  |  |  |  |
|  | Reoccurrence of illness | -0.275 | -3.38 | <0.001 |  |  |  |  |
|  | Age | -0.260 | -3.09 | 0.002 |  |  |  |  |
|  | Gender | -0.240 | -2.99 | 0.003 |  |  |  |  |
|  | Z WC + z BMI | 0.202 | 2.44 | 0.016 |  |  |  |  |

WC: waist circumference, ACE: adverse childhood experiences, CIRS: compensatory immunoregulatory system

### Negative acute phase proteins

**Table 4** shows that transferrin, albumin, and the z albumin + z Transferrin index were significantly correlated with HAMD, HAMA, FF, and STAI scores and with ROI. In addition, albumin, and the z albumin + z transferrin index were associated with total ACEs and the CIRS immune profile. Models #4, #5, and #6 in Table 5 show the results of multiple regression analysis with the negative acute phase proteins as dependent variables. We found that 20.1% of the variance in serum albumin was explained by weight loss, total ACEs, and the CIRS profile (all inversely) and male sex. After entering ROI, we found a somewhat better prediction whereby 21.8% of the variance was explained by ROI and weight loss (both inversely) and male sex. The same table, model #6 shows that serum transferrin was best explained by ROI, weight loss, age, and female gender (all inversely associated).

### Prediction of the rating scale scores

**Table 6** shows the prediction of the rating scale scores using metabolic, immunological, and demographic data as explanatory variables. Model #1 shows that 31.5% of the variance in the HAMD score was explained by total ACEs, CIRS profile, male sex (all positively), and the z albumin + z transferrin index. We found that 22.4% of the variance in the total HAMA score was explained by total ACEs (positively) and albumin (inversely). Up to 28.2% of the variance in the FF score was predicted by total ACEs and male gender (positively) and serum albumin (inversely). A large part of the variance in the total STAI score (33.3%) was explained by total ACEs (positively) and transferrin (inversely). We found that 35.4% of the variance in the integrated clinical severity index (ROI was not entered in this regression) was explained by total ACEs and male gender (positively) and transferrin and albumin (both inversely). Entering ROI in the regression analysis, showed that 53.0% of the variance in the integrated severity index was explained by ROI, total ACEs, male gender (positively), and albumin (inversely). This table also shows the results of regression analyses that examine the associations of ROI with ACEs and biomarkers. Model #7 shows that 35.3% of the variance in ROI was associated with total ACEs (positively) and z albumin + z Transferrin (inversely).

**Table 6.** Results of multiple regression analysis with the severity rating scales as dependent variables and biomarkers as explanatory variables.

| Dependent Variables | Explanatory Variables | Coefficient statistics |  |  | Model statistics |  |  |  |
| --- | --- | --- | --- | --- | --- | --- | --- | --- |
| | | $\beta$ | t | p | R<sup>2</sup> | F | df | p |
| #1. Total HAMD | **Model** |  |  |  | 0.315 | 17.57 | 4/153 | <0.001 |
|  | z Alb + z Tf index | -0.300 | -4.31 | <.001 |  |  |  |  |
|  | Total ACEs | 0.368 | 5.16 | <.001 |  |  |  |  |
|  | CIRS profile | 0.190 | 2.79 | 0.006 |  |  |  |  |
|  | Gender | 0.176 | 2.52 | 0.013 |  |  |  |  |
| #2. Total HAMA | **Model** |  |  |  | 0.224 | 22.37 | 2/155 | <0.001 |
|  | Total ACEs | 0.320 | 4.36 | <0.001 |  |  |  |  |
|  | Albumin | -0.273 | -3.72 | <0.001 |  |  |  |  |
| #3. Total FF | **Model** |  |  |  | 0.282 | 20.83 | 3/159 | <0.001 |
|  | Total ACEs | 0.428 | 5.95 | <0.001 |  |  |  |  |
|  | Albumin | -0.270 | -3.81 | <0.001 |  |  |  |  |
|  | Gender | 0.162 | 2.27 | 0.025 |  |  |  |  |
| #4. Total STAI, state | **Model** |  |  |  | 0.333 | 36.42 | 2/146 | <0.001 |
|  | Total ACEs | 0.525 | 7.75 | <.001 |  |  |  |  |
|  | Transferrin | -0.210 | -3.11 | 0.002 |  |  |  |  |
| #5. PC HAMD, HAMA, STAI, FF | **Model** |  |  |  | 0.354 | 20.27 | 4/148 | <0.001 |
|  | Total ACEs | 0.464 | 6.52 | <0.001 |  |  |  |  |
|  | Transferrin | -0.133 | -1.81 | 0.073 |  |  |  |  |
|  | Albumin | -0.247 | -3.20 | 0.002 |  |  |  |  |
|  | Gender | 0.156 | 2.11 | 0.037 |  |  |  |  |
| #6. PC HAMD, HAMA, STAI, FF | **Model** |  |  |  | 0.530 | 40.65 | 4/144 | <0.001 |
|  | Reoccurrence of illness | 0.535 | 7.61 | <0.001 |  |  |  |  |
|  | Albumin | -0.200 | -3.18 | 0.002 |  |  |  |  |
|  | Gender | 0.168 | 2.72 | 0.007 |  |  |  |  |
|  | Total ACEs | 0.190 | 2.68 | 0.008 |  |  |  |  |
| #7. Reoccurrence of illness | **Model** |  |  |  | 0.353 | 39.88 | 2/146 | <0.001 |
|  | Total ACEs | 0.506 | 7.47 | <0.001 |  |  |  |  |
|  | Z Alb + z Tf | -0.230 | -3.39 | <0.001 |  |  |  |  |

Alb: albumin, Tf: transferrin, ACE: adverse childhood experiences, CIRS: compensatory immunoregulatory system

### Results of PLS analysis

**Figure 1** illustrates the final PLS path model exclusively presenting significant paths (except from hsCRP to the phenome) and indicators. CTA demonstrated that the latent vectors were not erroneously represented as reflective models. We observed satisfactory convergence and construct reliability validity values for the phenome latent vector, with AVE = 0.821, Cronbach’s alpha = 0.927, rho A = 0.927, and composite reliability = 0.948. We observed satisfactory convergence and construct reliability validity values for the ACE latent vector, with AVE = 0.610, Cronbach’s alpha = 0.780, rho A = 0.840, and composite reliability = 0.861. We observed satisfactory convergence and construct reliability validity values for the IRS/CIRS latent vector, with AVE = 0.867, Cronbach’s alpha = 0.847, rho A = 0.851, and composite reliability = 0.929. The model’s overall quality is satisfactory, evidenced by an SRMR of 0.034. We found that 51.3% of the variance in the depression phenome was explained by z albumin + z transferrin, ROI, ACEs, and CIRS, whilst hsCRP was not significant. Up to 17.1% of the variance in the negative APP response was explained by age, IRS/CIRS, ROI, and BMI. A large part of the variance in hsCRP (25.3%) was explained by ACEs, M1 macrophage, CIRS profile, waist circumference, BMI, and sex. The effects of ROI on the phenome are in part mediated by z albumin + z transferrin (t=0.012). Female sex (sex is entered as male 1, female = 0) impacts the phenome and its effects are mediated by ACEs (t=-3.41, p<0.001), the path from ACEs to ROI (t=-3.20, p=0.001), and the path from ACEs to ROI to z albumin + z transferrin (t=-1.72, p=0.043). The analysis of total effects showed significant effects of ACEs (t=7.91, p<0.001), age (t=1.95, p=0.025), BMI (t=1.75, p=0.041), CIRS (t=2.15, p=0.016), IRS/CIRS (t=1.85, p=0.032), ROI (t=9.26, p<0.001), female sex (t=3.41, p<0.001), and z albumin + z transferrin (t=-2.79, p=0.003) on the phenome.

**Figure 1.**
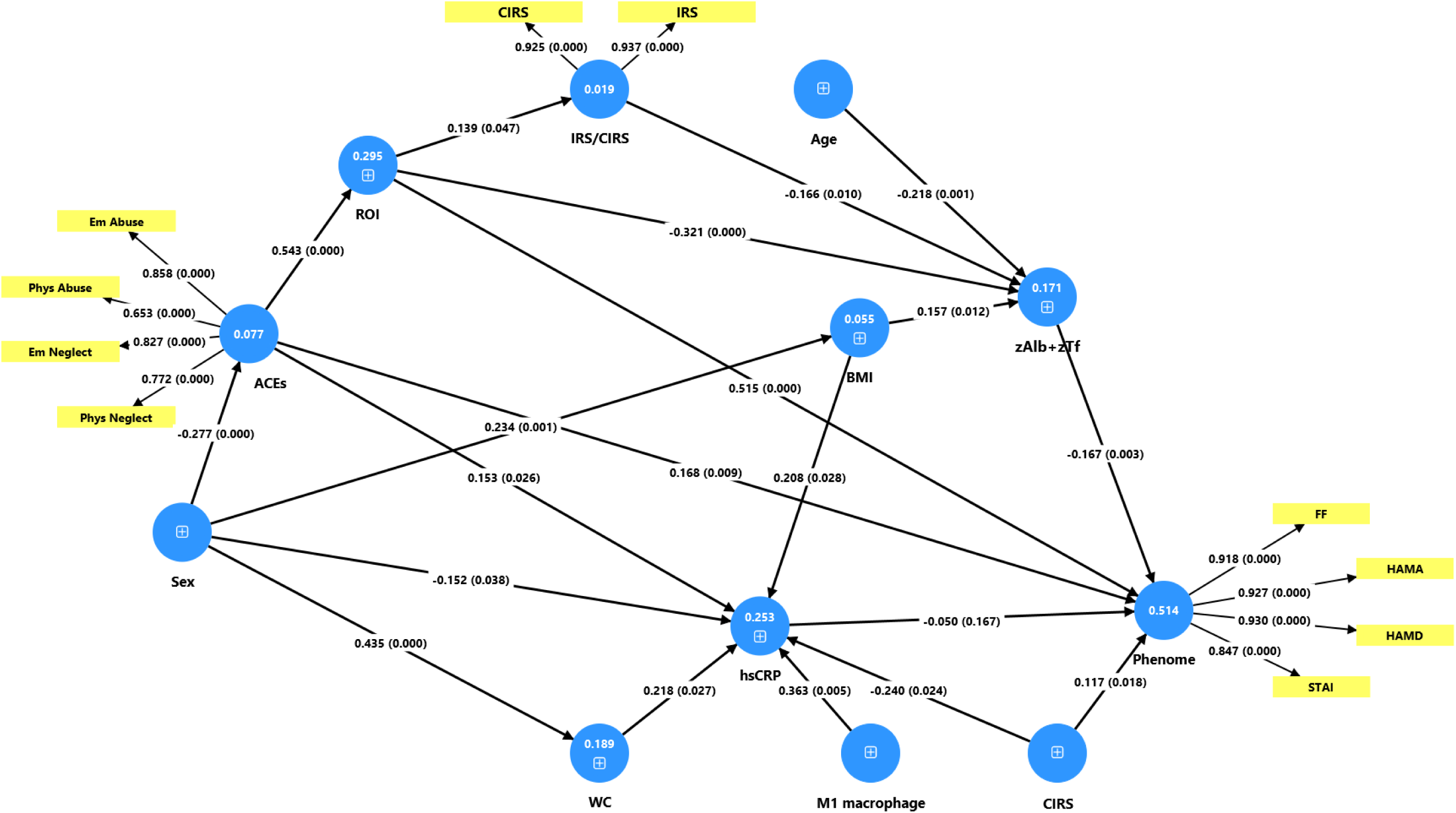
Results of partial least squares analysis (PLS) showing the causal pathways between explanatory latent vectors or single indicators and the final outcome variable, namely a latent vector of the physio-affective phenome of major depression. The latter was extracted from four rating scale scores, namely FF: Fibro-Fatigue scale; HAMD: Hamilton Depression Rating Scale; HAMA: Hamilton Anxiety Rating Scale; and STAI: State-Trait Anxiety Inventory. This model shows the pathway coefficients (with p values) and the loadings (p-value) of the latent vectors. The figures in the blue circles denote the explained variance. z Alb + z Tf: sum of z scores of albumin + transferrin; IRS/CIRS: Immune-inflammatory response system / Compensatory immunoregulatory system; ROI: reoccurrence of illness index; ACE: adverse childhood experiences; BMI: body mass index; hsCRP: high-sensitivity C-reactive protein; WC: waist circumference; sex: binary variable with male=1, female=0.

## Discussion

### Serum hsCRP is not increased in MDD

The first major finding of this study is that there are no significant elevations in serum hsCRP levels in individuals with MDD compared to controls. This study identified, using multivariate statistical analyses, even an inverse correlation between hsCRP and MDD and the severity of depression. We previously reported that serum CRP concentrations measured with the standard assay are not elevated in MDD [34]. Certain authors employed a cut-off threshold of 6 mg/dL to exclude individuals exhibiting overt inflammation while still identifying alterations in IRS and CIRS cytokine profiles in MDD [35]. A blunted hsCRP response does not preclude the presence of an inflammatory response as this phenomenon might be noted even in severe inflammatory or autoimmune conditions such as systemic lupus erythematosus, scleroderma, polymyositis, and certain viral infections [36–39].

Meta-analyses utilizing the hsCRP assay indicated elevated blood levels in MDD relative to controls [40–43]. The data from all those meta-analyses were interpreted to suggest a correlation between MDD and inflammation. Consequently, certain authors employ serum hsCRP levels > 3 mg/L as a benchmark to indicate “inflammatory depression” or “immune-mediated depression” [11]. According to this criterion, around 25-30% of people with MDD would experience inflammation. Since only 25%-30% of MDD individuals may exhibit elevated hsCRP levels, it remains uncertain if the mean hsCRP levels are always elevated in study samples with that disorder. In our study, the majority of MDD patients (84.8%) had hsCRP values below 3 mg/L, whereas only 15.2% demonstrated values over 3 mg/L. According to our introduction, serum concentrations of 3 to 10 mg/L indicate normal or slightly raised levels, while values between 10 mg/L and 100 mg/L signify significant inflammation, as seen in many autoimmune illnesses [18]. In the present investigation, we observed that 15 of the 19 patients with elevated hsCRP levels exhibited values ranging from 3 to 7 mg/L, whereas only 4 individuals demonstrated levels between 30 and 40 mg/L. Consequently, merely 4 out of the 125 MDD patients (3.2%) exhibit values that could suggest moderate IRS activation. Rephrased, the majority of MDD patients in our study exhibit hsCRP values that do not suggest overt inflammation. Consequently, it is erroneous to attribute minor elevations in hsCRP as indicators of “inflammatory depression,” particularly given the multitude of metabolic disorders relevant to MDD that correlate with heightened hsCRP, which will be elaborated upon in the subsequent section.

### hsCRP is a marker of metabolic alterations, ACEs, and immune homeostasis

The second major finding of this study is that increased serum hsCRP in the lower concentration ranges is largely associated with metabolic parameters including MetS, the ranking of MetS, BMI and waist circumference. These associations were detected when examining the total study group, but also in the restricted study groups of individuals with and without increased hsCRP values using a threshold of 3 mg/L. In those three study samples, metabolic variables were the most important determinants of increased hsCRP values. This is in agreement with the knowledge that increased hsCRP in the low concentration range (<100 mg/L) reflects in part metabolic changes including BMI, and metabolic conditions or disorders (see introduction), and that values >13 mg/L reflect a special phenotype of MetS, characterized by IRS activation, increased uric acid, platelet activation and liver dysfunction [18]. Moreover, hsCRP levels in the low concentration range are used as a act as a prognostic indicator for cardiovascular disease and atherosclerosis when exceeding > 3 mg/L [14–18, 44–47].

In this regard, it is crucial to acknowledge that the majority of the meta-analyses on CRP in MDD, as discussed in the previous section, did not account for BMI or MetS. For instance, Dowlati et al (2010) and Kohler et al. (2014) did not consider the hsCRP data for BMI or MetS [40, 41]. In the other meta-analyses mentioned, not all included studies could be controlled for BMI or MetS. In addition, all of these studies were unable to account for the ranking of MetS, waist circumference, or the numerous metabolic conditions (see Introduction) that are associated with elevated hsCRP. Consequently, it is evident from our regression and PLS analyses that the results of hsCRP measurements in MDD will be contingent upon the number of patients enrolled in the study who have an increased BMI, waist circumference, MetS, or obesity. This situation is further complicated by the comorbidity between MDD and MetS and the considerable interactions between MetS and atherogenicity and immune characteristics, which result in an increased severity of depression [1, 48]. In reality, we demonstrated that certain biomarkers, such as lipids and immune data, cannot be assessed in depression without excluding patients with MetS or strictly controlling for MetS [48, 49]. This is of paramount importance, as the majority of studies in MDD fail to account for MetS, obesity, elevated BMI, or other metabolic conditions even when they investigate lipids and atherogenicity [49].

Furthermore, we observed that the most accurate prediction of hsCRP was achieved by incorporating the cumulative effects of BMI, waist circumference, ACEs, the M1 macrophage and CIRS profiles in multiple regression or PLS analysis. Moraes et al. [21] reported that a significant portion of the variance in hsCRP (approximately 50%) was accounted for by BMI, sex, and early childhood sexual abuse. Prior to this, there were reports that ACEs could result in elevated CRP levels in later life [50–54]. Interestingly, ACEs may also be linked to an elevated incidence of MetS later in life [55, 56].

The present investigation also demonstrates that the activation of M1 macrophages contributes to the elevated levels of hsCRP. This is consistent with the understanding that M1 proinflammatory cytokines can trigger the APP response in the liver, thereby resulting in the production of positive APPs [3]. Conversely, we discovered that the production of hsCRP is downregulated by increased CIRS activity. It is well-established that cytokines, including IL-4 and IL-10, influence the AP response in the liver by decreasing the expression of APPs in response to pro-inflammatory cytokines and suppressing IL-6 production [57, 58]. In addition, EGF may inhibit the production of APPs by decreasing IL-6 and TNF-α, as well as protecting against hepatic inflammation [59, 60].

Based on the above, it is reasonable to infer that the levels of hsCRP in the low concentration range in MDD without overt inflammation or infection are indicative of metabolic health, the impact of early childhood experiences, and the IRS-CIRS equilibrium in MDD. As previously reviewed [1], MDD is distinguished by elevated IRS and CIRS activities, with the IRS system frequently being the dominant system during the acute phase of the illness [5]. However, in certain phases of MDD (e.g., partial remission), the CIRS can be more dominant [1]. Therefore, one hypothesis is that the homeostasis setpoint between IRS and CIRS may be reflected in hsCRP levels after the effects of metabolic data and metabolic conditions have been accounted for. Additionally, it is imperative to emphasize that the IRS/CIRS equilibrium is different among MDD phenotypes, which encompasses simple dysmood disorder and major dysmood disorder, in addition to the phase of the index episode (acute, remission, or partial remission) [1]. By inference, all those factors may influence serum hsCRP concentrations.

### The negative AP response versus hsCRP

The third significant discovery of this study is that, although elevated hsCRP cannot be regarded as a biomarker of MDD, serum transferrin and albumin levels are significantly reduced in MDD and are strongly correlated with the affective (depression and anxiety) and physiosomatic (fibro-fatigue symptoms) phenome of MDD. The results obtained in the current study in Chinese patients are consistent with those of our research publications, which were published at various institutions in Europe between 1991 and 1997 [2, 4]. There are currently some studies that demonstrate a decrease in albumin and transferrin levels in patients with MDD [61–63].

As previously mentioned in the 1990s, this data suggests that MDD is accompanied by a negative acute phase response and protein deficiency or disruptions in protein homeostasis [2, 34]. The latter discovery may be in part the result of primary undernutrition caused by anorexia and weight loss, which are critical symptoms of MDD [2]. Weight loss and IRS/CIRS activation were significant determinants of decreased transferrin and albumin levels in the present study. Therefore, it is possible that protein malnutrition as a result of anorexia and weight loss results - in conjunction with IRS activation - in decreased levels of both visceral proteins. In fact, the M1 cytokines elicit anorexia and weight loss, which could ultimately lead to protein metabolism disorders [3]. In the AP response, the body prioritizes “immune functions” over the synthesis of low-priority proteins (such as albumin), resulting in the conversion of albumin to energy sources and the production of positive APPs [3].

Consequently, our data set demonstrates a comprehensive picture of non-classic inflammation, in which a positive acute phase protein, such as hsCRP, is not elevated and may even be reduced in MDD, while the negative APPs are downregulated. This condition may suggest a smoldering low-grade inflammation, chronic undernutrition, increased catabolism, nutritional deficiency, and impaired hepatic synthetic function [64–66].

It is important to recognize that the negative AP response in MDD is influenced by ACEs. The inverse effect of ACEs on these visceral proteins contributes to the extensive body of literature that demonstrates the impact of ACEs on a variety of NIMETOX pathways in MDD, such as atherogenicity, IRS and CIRS, the gut microbiome, oxidative stress, and antioxidant levels [1].

As described in the Introduction, some authors maintain that the prevalence of “inflammatory depression” is approximately 25-30%, as determined by the hsCRP criterion, and that these patients would respond favorably to anti-inflammatory medication treatment [11]. This logic implies that the remaining 60-70% would not exhibit “inflammatory depression” or “immune-mediated depression.” However, our investigation demonstrated that a combination of reduced APPs has an external validating capacity for MDD, as evidenced by an AUC of 0.778. Consequently, the combination of negative APPs was significant, whereas hsCRP did not produce any significant diagnostic performance for MDD. In the past, we discovered that this combination was highly accurate for MDD, particularly melancholia, with a sensitivity of 72% and a specificity of 92% [2]. Moreover, the AUC was further enhanced to 0.798 by combining both visceral proteins, the CIRS profile, and IL-17A. We demonstrated already in the 1990s that the combination of various immune and APP biomarkers resulted in an adequate diagnostic performance for MDD [4]. Recently, we demonstrated that the combination of various NIMETOX pathways indicates that approximately 78.8% of MDD patients exhibit aberrations in those pathways [48]. Our data indicate that a significant number of patients with MDD, if not all, manifest different and occasionally divergent aberrations in NIMETOX pathways [1, 48].

### Conclusions

hsCRP is not a valid biomarker of MDD. Metabolic variables significantly influence this positive APP. MDD is characterized by decreased levels of the negative APPs albumin and transferrin. The Th-17 and CIRS immune profiles together with these negative APPs enable the identification of approximately 65% of MDD patients with a reasonable degree of specificity. Therefore, assertion that “inflammatory depression” would be present in 25-30% of MDD patients is unfounded and should be replaced by the argument that many, if not all, MDD patients exhibit NIMETOX anomalies, including a negative AP response. The latter suggests protein malnutrition and non-classical inflammation. The negative acute phase response should be considered as a novel treatment target in MDD. This can be achieved by a) addressing the underlying immune activation; b) providing nutritional support, particularly protein supplementation; and c) supporting hepatic protein synthesis.

## Data Availability

The first author (MM) will provide access to the dataset supporting this study upon receipt of a valid request and the completion of a thorough data review.

## Acknowledgements

Not Applicable

## Ethical approval and consent to participatg

The study was approved by the Research Ethics Committee of the Sichuan Provincial People’s Hospital, Chengdu, China [Ethics (Research) 2024-203]. Written informed consent was obtained from all participants.

## Declaration of interest

No competing of interest to be declared by authors.

## Funding

This research was funded by the Health Science Research Project of Sichuan Province (Grant No.: ZH2024-203) and Sichuan Science and Technology Program “PIANJI” Project (Grant No.: 2025HJPJ0004).

## Author’s contributions

All authors contributed equally to this research and approved the final version of the paper.

## Notes

### Competing Interest Statement

The authors have declared no competing interest.

